# Relationship between cognition and frailty-A cross-sectional study among elders in the tribal areas of Mysuru

**DOI:** 10.1101/2023.01.31.23285235

**Authors:** A Vanmathi, Praveen Kulkarni, Dennis Chauhan, D Sunil Kumar, Prathiba Periera, M Kishor, Sayana basheer

## Abstract

**Background:** Cognition declines as age advances. Frailty is a pre-disability state. cognitive impairment and frailty lower the quality of life of elders. studies done in this background were mostly in urban or rural areas. Hence the current study was conducted among elders in the tribal areas to find the relationship between cognition and frailty.

**Materials and methods:** It was a cross-sectional study done for a period of one year among 316 elders >60 years. Based on the reported prevalence of cognitive dysfunction among the elderly to be 25% (Rakesh M Patel et al), with 5% absolute allowable error,5% of alpha error, and 10% non-response rate) residing in the tribal areas of Mysuru. WHO-30 cluster sampling was used. MOCA-B was used for assessing cognition scale and the TFI scale was used for assessing frailty.

**Results:** The prevalence of cognitive impairment and frailty was 95.3 % and 61.4% respectively. Factors like Gender, education, type of family, physical, financial and emotional dependency, and preference of treatment were associated with cognition. There was also a significant association between cognition and frailty.

**Conclusion:** There was a higher prevalence of cognitive impairment among frail elders. Thus, early diagnosis and appropriate management are necessary for healthy ageing

## Introduction

Ageing gracefully is an art ^[1].^

According to WHO, one in six individuals in the world will be 60 years of age or older by the year 2030. By 2050, 80 per cent of elderly citizens will reside in developing and middle-income nations ^[2]^ In India there were about 104 million elders according to 2011 census ^[3]^

Ageing is a biological process caused by many cellular and molecular damages resulting in the deterioration of physical and mental abilities leading to increased susceptibility to diseases and eventually death. ^[4]^ While some changes in older people’s health are inherited, the majority are caused by people’s living conditions, neighbourhoods, and communities, as well as by their characteristics such as their sex, race, or financial position. ^[2]^

Tribals constitute about 8.6% of the total Indian population. People in the tribal region are the most neglected and vulnerable population with a high degree of malnutrition, morbidity and mortality. The tribal elders play the role of culture transformer and this transformation predisposes them to psychosocial issues such as depression, anxiety, stress, and cognitive deficiencies ^[5],[6]^

Cognitive function is one aspect of overall brain function and is an essential component for carrying out daily tasks ^[7]^. Cognitive dysfunction in the elderly is characterised by loss of memory, Asking the same question repeatedly or telling the same tale repeatedly, Losing sight of familiar faces and surroundings, and Experiencing troubles in judgement, such as being unable to decide what to do in an emergency, Modifications in behaviour or mood, eyesight issues, Having trouble organising and completing tasks, like following a recipe or keeping track of monthly bills Furthermore, it has substantial social effects, resulting in a loss of autonomy and independence, as well as increasing demand for permanent caregivers and healthcare help. ^[8]^

Poor nutrition, Midlife obesity, hyperlipidemia, midlife hypertension, diabetes, smoking, alcohol use, sleep disturbances, and depression are among the modifiable risk factors for cognitive dysfunction. While other factors, such as physical activity, education, and social engagement, reduce the risk of cognitive dysfunction. The development of cognitive impairment lowers the quality of life, leading to severe painful physical and emotional stress. ^[9],[10]^Frailty is a pre-disability state American Geriatrics Society describes frailty as *“a state of increased vulnerability to stressors due to age-related declines in physiologic reserve across neuromuscular, metabolic, and immune systems”* ^[11]^. Frailty May lead to several adverse health outcomes such as falls, disability, hospitalization ^[12]^ etc, recently it has been linked to cognition also Most of the reported studies in this field have been conducted in hospitals or on older people living in rural or urban areas. Despite the fact that few studies have been conducted in tribal regions, they are related to general health conditions among the elderly. Taking this into consideration, we propose the current study to find the relationship between nutrition and cognitive functioning of the elderly living in tribal areas of Mysuru.

## Method and materials

A cross-sectional study was done for a period of 6 months among elders >60 years residing in the tribal areas of Mysuru for a period of 6 months. A total of 316 elders (Based on the reported prevalence of cognitive dysfunction among the elderly to be 25% (Rakesh M Patel et al) ^[13]^, with 5% absolute allowable error and 5% of alpha error and 10% non-response rate) who were willing to participate in the study were included after obtaining necessary consent. In case, the elderly was illiterate/not able to comprehend, consent was obtained in presence of an impartial witness. Elders who had visual and auditory deficiencies, those who were not able to speak and diagnosed with mental illness were excluded

WHO-30 clusters sampling technique ^[14]^ was used The line listing of the population of all the tribal hamlets was done by collecting data from local panchayats and the cumulative total was calculated. The cumulative population was divided by sample size (316) to get the sampling interval. In each cluster total of 11 elders were interviewed (316/30). The first cluster was identified from the list where the sampling interval exists and the subsequent cluster was identified by adding the sampling interval to the population of the previous cluster. During the survey, the centre of the hamlet was identified and one road was randomly selected. A household survey was conducted on the elders satisfying the inclusion and exclusion criteria who were residing in the house. If a household had more than one elderly, only one was chosen randomly irrespective of gender. If a house was locked, an elderly from the subsequent house was interviewed. If a hamlet didn’t have enough samples (i.e.<11 elders), elders from other hamlets which had a higher elderly population were chosen. Ethical committee approval was obtained before data collection.

The data collection had three parts. a) sociodemographic profile, b) cognitive assessment using MOCA -B scale ^[15]^. c)Tilburg frailty indicator ^[16]^(TFI) for assessing frailty

The data thus obtained was coded and entered into MS excel 2019 spreadsheet and analysed using SPSS version 25 (licensed to JSSAHER). Data were expressed using descriptive statistics like frequency, percentage and proportion. The Chi-square test/Fisher’s Exact Test was used to find the association between various socio-demographic variables and cognition and also with frailty. A correlation between frailty and cognition was found using the Pearson correlation. A p-value of < 0.05 was considered statistically significant.

## Results

### Sociodemographic profile

In the study comprising 316 elderlies, most of the participants were females (60.8%,192) And the majority belonged to the age group of 60-65 years (56.6%, 179) and 25.3% (80) were in the age group of 66-70 years. Most of the elderly interviewed were illiterates (82.3%,260) and very few were literates (17.7%,56). only 46.8%,148 were currently working whereas 53.2%, (168) were not involved in any occupation. Almost 99.1%,313 were Hindus. More than half of the elders were married (58.2%,184),36.4 %,115 were widowers 16,5.1% were unmarried. 70.9%, (224) belonged to the nuclear family and27.8%,88 were in a joint family,

Among the study participants 63%,199 stated they were not dependent on others for their day-to-day activities and 4,1.3% were fully dependent on others, 55.4% (175) participants were independent financially while 15,4.7% were completely dependent on others When questioned about their emotional dependency 101,32 % stated that they were fully involved in family decisions whereas 153,48.4% were partially involved and 62,19.6% stated that they were never involved

Out of 316 elders interviewed 39(12.3%) had comorbidities. More than half of the study population (59.5%,188) preferred allopathy as their first preference,34.5%,109 preferred home remedies, 55.7% (176) had addictive habits and 32.3%, 102 said that they have sleep disturbances (Table 1)

**Table 1:** socio-demographic profile of the study population.

| **Variables** | **Categories** | **Frequency** | **Percent (%)** |
| --- | --- | --- | --- |
| **Gender** | Male | 124 | 39.2 |
|  | Female | 192 | 60.8 |
| **Age** | 60-65 | 179 | 56.6 |
|  | 66-70 | 80 | 25.3 |
|  | 71-75 | 34 | 10.8 |
|  | >75 | 23 | 7.3 |
| **Education** | Literate | 56 | 17.7 |
|  | Illiterate | 260 | 82.3 |
| **Currently working** | Yes | 148 | 46.8 |
|  | No | 168 | 53.2 |
| **Religion** | Hindu | 313 | 99.1 |
|  | Muslim | 2 | 0.6 |
|  | Christian | 1 | 0.3 |
| **Marital status** | Married | 184 | 58.2 |
|  | Widow/Widower | 115 | 36.4 |
|  | Separated | 1 | 0.3 |
|  | Unmarried | 16 | 5.1 |
| **Type of family** | Nuclear | 224 | 70.9 |
|  | Three generation | 4 | 1.3 |
|  | Joint/extended | 88 | 27.8 |
| **Living status** | Spouse and children | 45 | 14.2 |
|  | Spouse | 68 | 21.5 |
|  | Children | 162 | 51.3 |
|  | Alone | 41 | 13 |
| **Physical dependency** | Independent | 199 | 63 |
|  | Partially dependent | 113 | 35.7 |
|  | Fully dependent | 4 | 1.3 |
| **Financial dependency** | Independent | 175 | 55.4 |
|  | Partially dependent | 126 | 39.9 |
|  | Fully dependent | 15 | 4.7 |
| **Emotional dependency** | Always | 101 | 32 |
|  | Sometimes | 153 | 48.4 |
|  | Never | 62 | 19.6 |
| **Co-morbidities** | Present | 39 | 12.3 |
|  | absent | 277 | 87.7 |
| **Treatment modalities** | Allopathy | 188 | 59.5 |
|  | Indigenous | 1 | 0.3 |
|  | Traditional | 8 | 2.5 |
|  | Home remedies | 109 | 34.5 |
|  | Magico-religious | 10 | 3.2 |
| **Addictive habits** | Present | 176 | 55.7 |
|  | Absent | 140 | 44.3 |
| **Sleep disturbances** | Present | 102 | 32.3 |
|  | Not present | 177 | 56 |
|  | Sometimes | 37 | 11.7 |

### Cognition

301,95.3% had cognitive impairment whereas 15, 4.7% were found to be normal (Fig 1), when graded (according to the moca B scale)13%,42 elders had severe cognitive impairment,54.1%,171 had moderate and 28.2%,89 had mild cognitive impairment (Fig 2)

**Fig 1:**
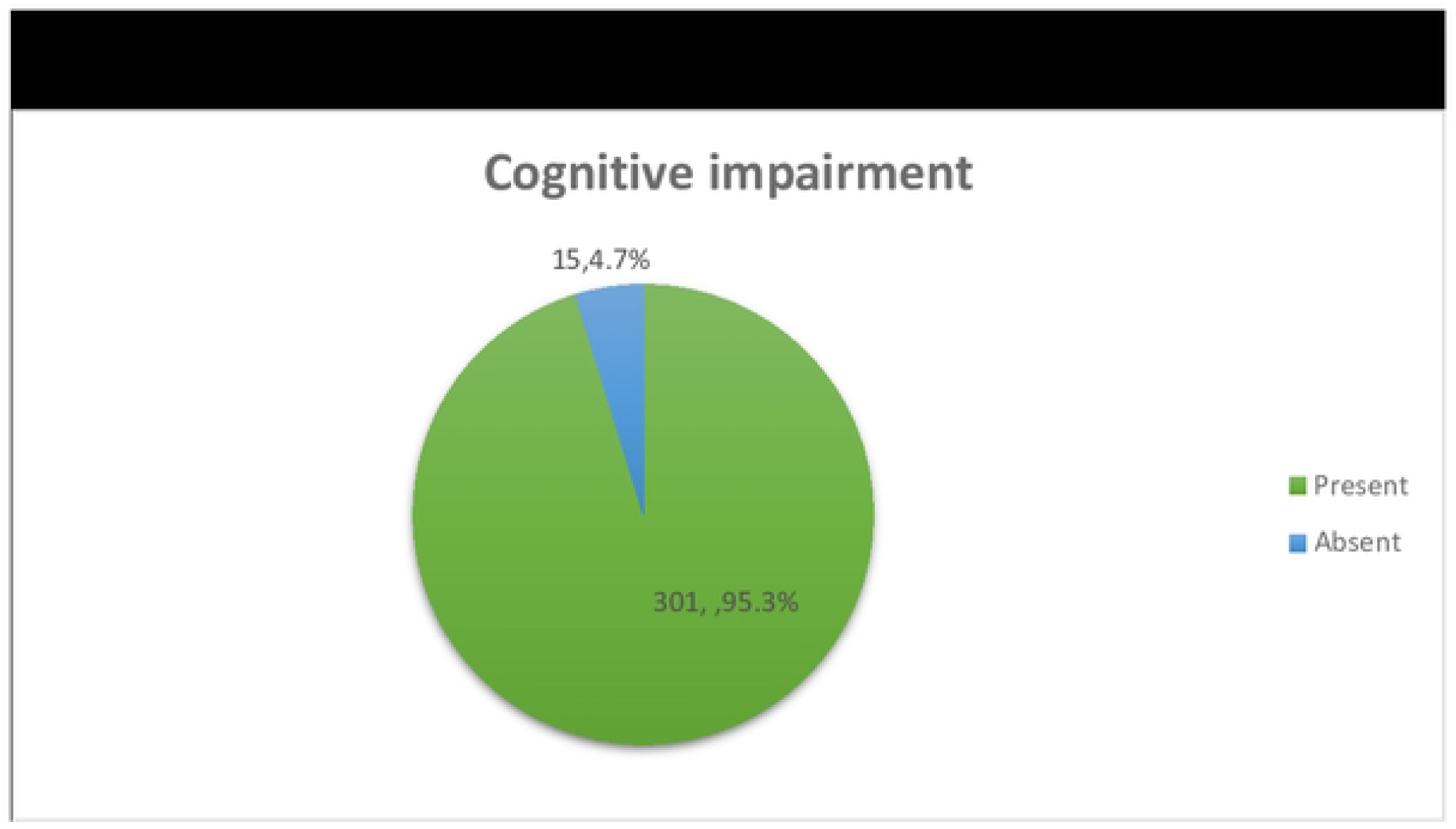
Prevalence of cognitive impairment among the study population.

**Fig 2:**
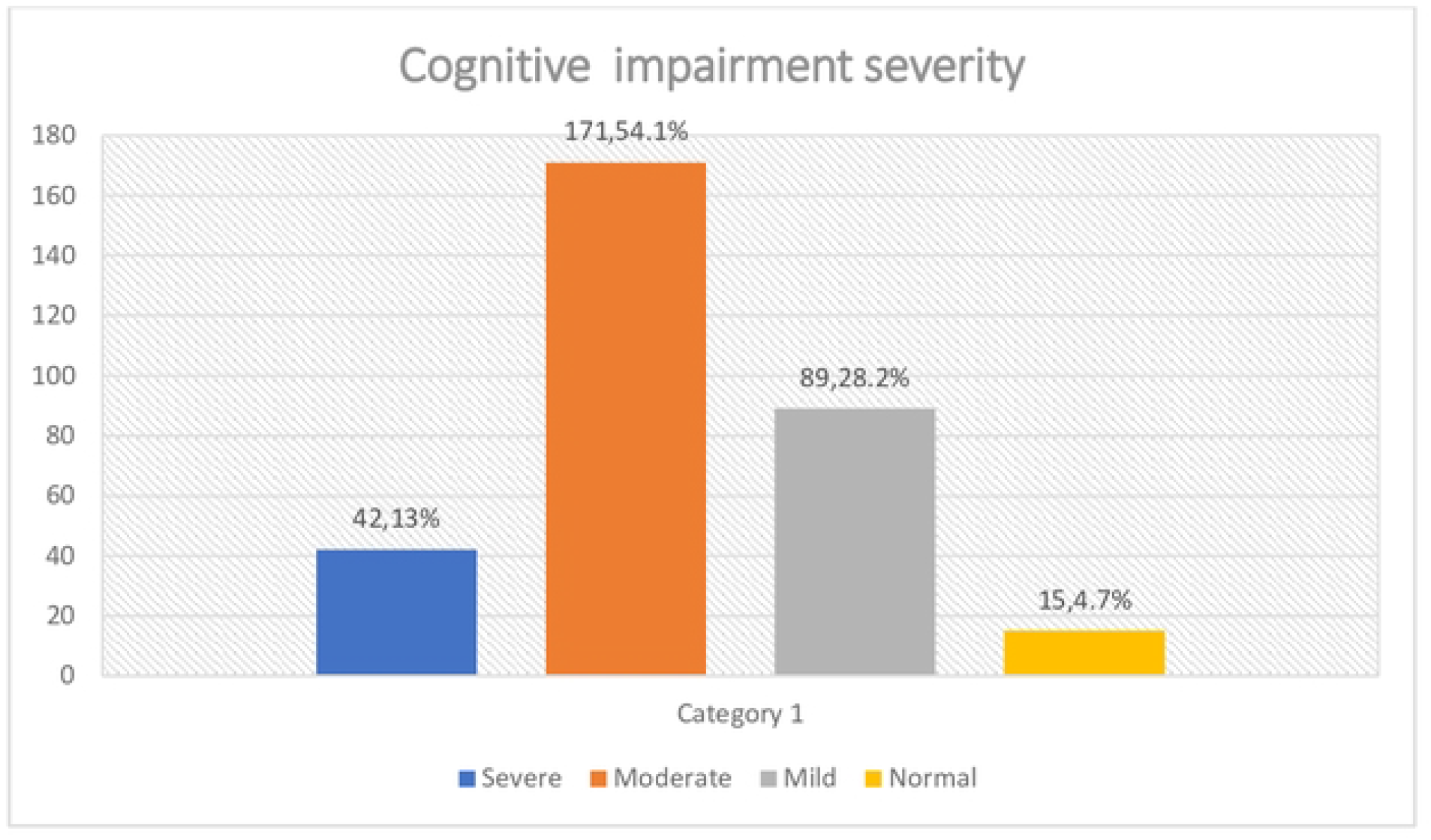
Distribution of study population according to the severity of cognitive impairment.

Gender, education, type of family, physical, financial and emotional dependency, and preference of treatment were associated with a cognitive impairment which was statistically significant (Table 2)

**Table 2:** Factors associated with cognitive impairment.

| **Variables** | **Categories** | **Cognitive impairment present** | **Cognitive impairment absent** | **Chi-Square value** | **P value** |
| --- | --- | --- | --- | --- | --- |

|  |  | **(301) (n,**<br>**%)** | **(15) (n,**<br>**%)** |  |  |
| --- | --- | --- | --- | --- | --- |
| **Gender** | Male | 113(37.5) | 11(73.3) | 7.677 | **0.006\*** |
|  | Female | 188(62.5) | 4(26.7) |  |  |
| **Age** | 60-65 | 167(55.5) | 12(80) | 2.560 | 0.409 |
|  | 66-70 | 78(25.9) | 2(13.3) |  |  |
|  | 71-75 | 33(11) | 1(6.7) |  |  |
|  | >75 | 23(7.6) | 0 |  |  |
| **Education** | Literate | 46(15.3) | 10(66.7) | 25.87<br>3 | **<0.001\*** |
|  | Illiterate | 255(84.7) | 5(33.3) |  |  |
| **Currently**<br>**working** | Yes | 138(45.8) | 10(66.7) | 2.487 | 0.115 |
|  | No | 163(54.2) | 5(33.2) |  |  |
| **Religion** | Hindu | 299(99.3) | 14(93.3) | 6.725 | 0.136 |
|  | Muslim | 1(0.3) | 1(6.7) |  |  |
|  | Christian | 1(0.3) | 0 |  |  |
| **Marital**<br>**status** | Married | 17(57.1) | 12(80.0) | 3.713 | 0.346 |
|  | Widow/Wido<br>wer | 112(37.2) | 3(20.0) |  |  |
|  | Separated | 1(0.3) | 0 |  |  |
|  | Unmarried | 16(5.3) | 0 |  |  |
| **Type of family** | Nuclear | 218(71.8) | 6(40.0) | 9.688 | **0.022\*** |
|  | Three generation | 3(1.0) | 1(6.7) |  |  |
|  | Joint/extended | 80(26.6) | 8(53.3) |  |  |
| **Living status** | Spouse and children | 42(14.0) | 3(20.0) | 2.720 | 0.450 |
|  | Spouse | 64(21.3) | 4(26.7) |  |  |
|  | Children | 154(51.2) | 8(53.3) |  |  |
|  | Alone | 41(13.6) | 0 |  |  |
| **Physical dependency** | Independent | 185(61.5) | 14(93.3) | 6.716 | **0.03\*** |
|  | Partially dependent | 112(37.2) | 1(6.7) |  |  |
|  | Fully dependent | 4(1.3) | 0 |  |  |
| **Financial dependency** | Independent | 161(53.5) | 14(93.3) | 8.926 | **0.011\*** |
|  | Partially dependent | 125(41.5) | 1(6.7) |  |  |
|  | Fully dependent | 15(5.0) | 0 |  |  |
| **Emotional**<br>**dependenc**<br>**y** | Always | 90(29.9) | 11(73.3) | 11.49<br>1 | **0.002\*** |
|  | Sometimes | 149(49.5) | 4(26.7) |  |  |
|  | Never | 62(20.6) | 0 |  |  |
| **Co-**<br>**morbidity** | Present | 36(12.0) | 3(20.0) | 0.854 | 0.410 |
|  | absent | 265(88.0) | 12(80.0) |  |  |
| **Treatment**<br>**modalities** | Allopathy | 179(59.5) | 9(60) | 9.193 | **0.05\*** |
|  | Indigenous | 0 | 1(6.7) |  |  |
|  | Traditional | 7(2.3) | 1(6.7) |  |  |
|  | Home<br>remedies | 105(34.9) | 4(26.7) |  |  |
|  | Magico-religious | 10(3.3) | 0 |  |  |
| **Addictive**<br>**habits** | Present | 168(55.8) | 8(53.3) | 0.036 | 1.000 |
|  | Absent | 133(44.2) | 7(46.7) |  |  |
| **Sleep**<br>**disturbanc**<br>**es** | Present | 95(31.6) | 7(46.7) | 3.792 | 0.102 |
|  | Not present | 172(57.1) | 5(33.3) |  |  |
|  | Sometimes | 34(11.3) | 3(20.0) |  |  |

n=Frequency,%=Percent,\*-Statistically significant p <0.05

### Frailty

61.4% (194) of the study population were frail according to the TFI scale whereas 38.6% (122) were non-frail (Fig 3).

**Fig 3:**
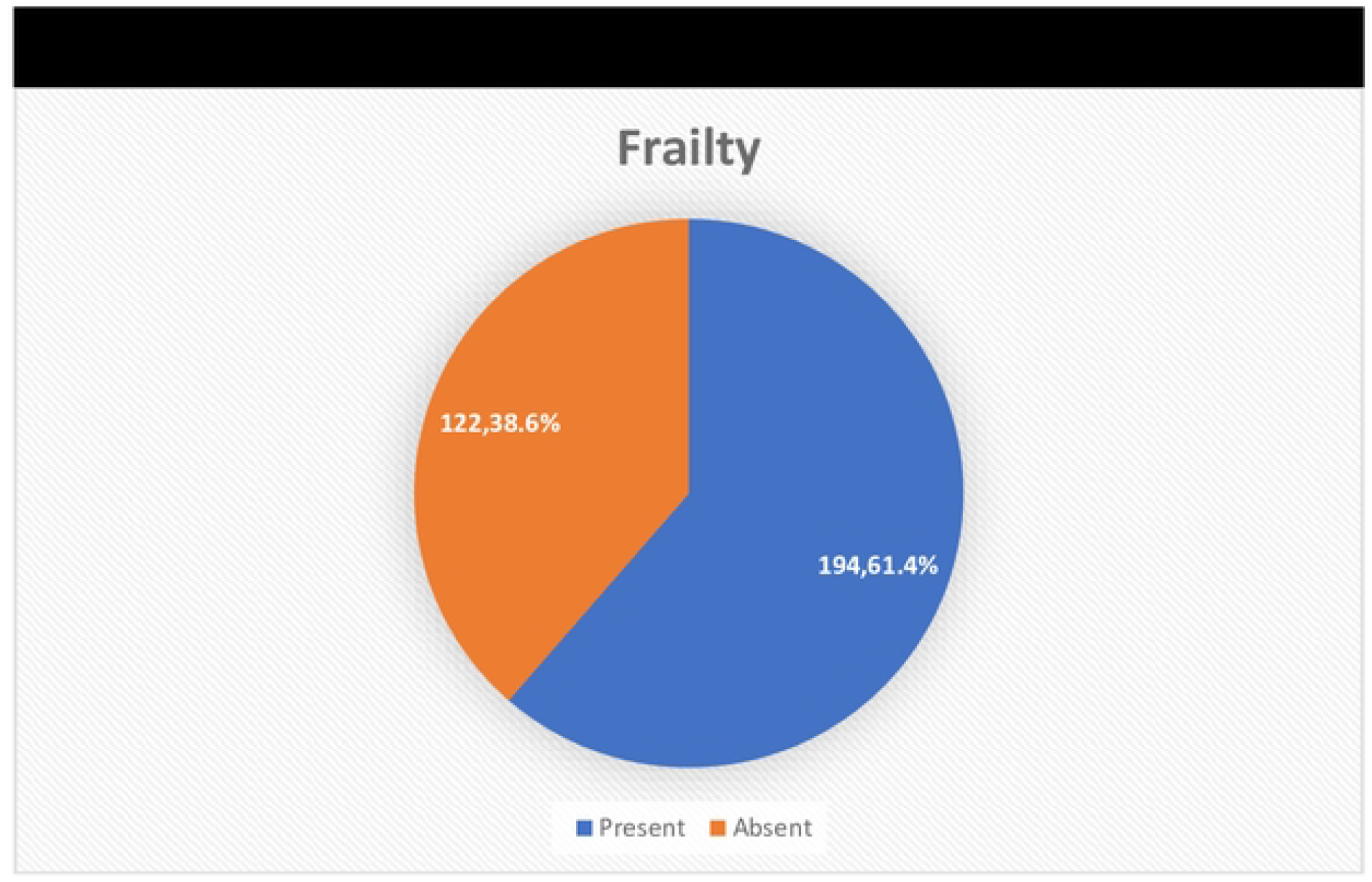
Distribution of study population based on frailty.

Factors like age, education, marital status, type of family, living status, Physical and emotional dependency and also the presence of comorbidities were significantly associated with frailty. (Table 3)

**Table 3:**
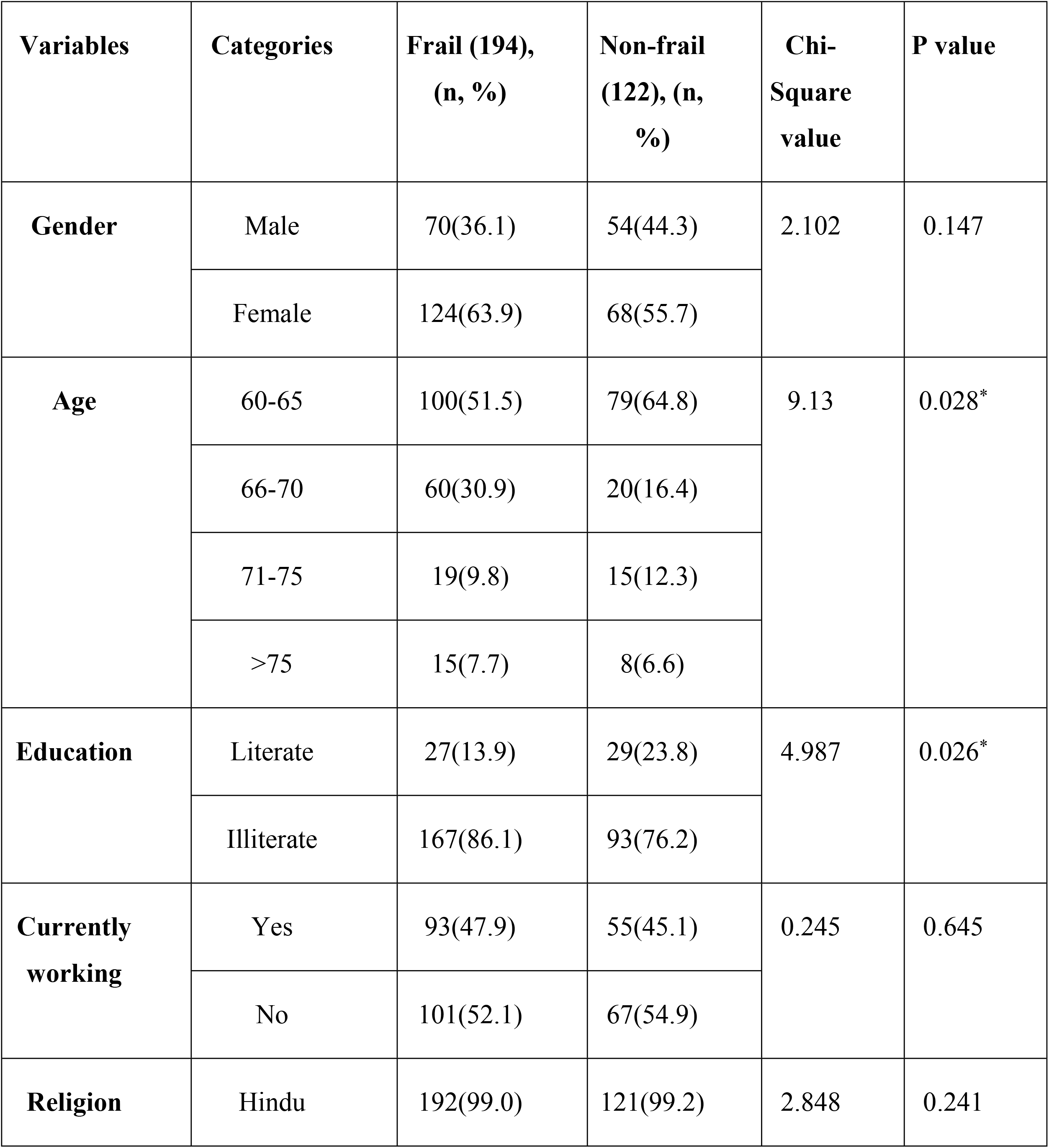
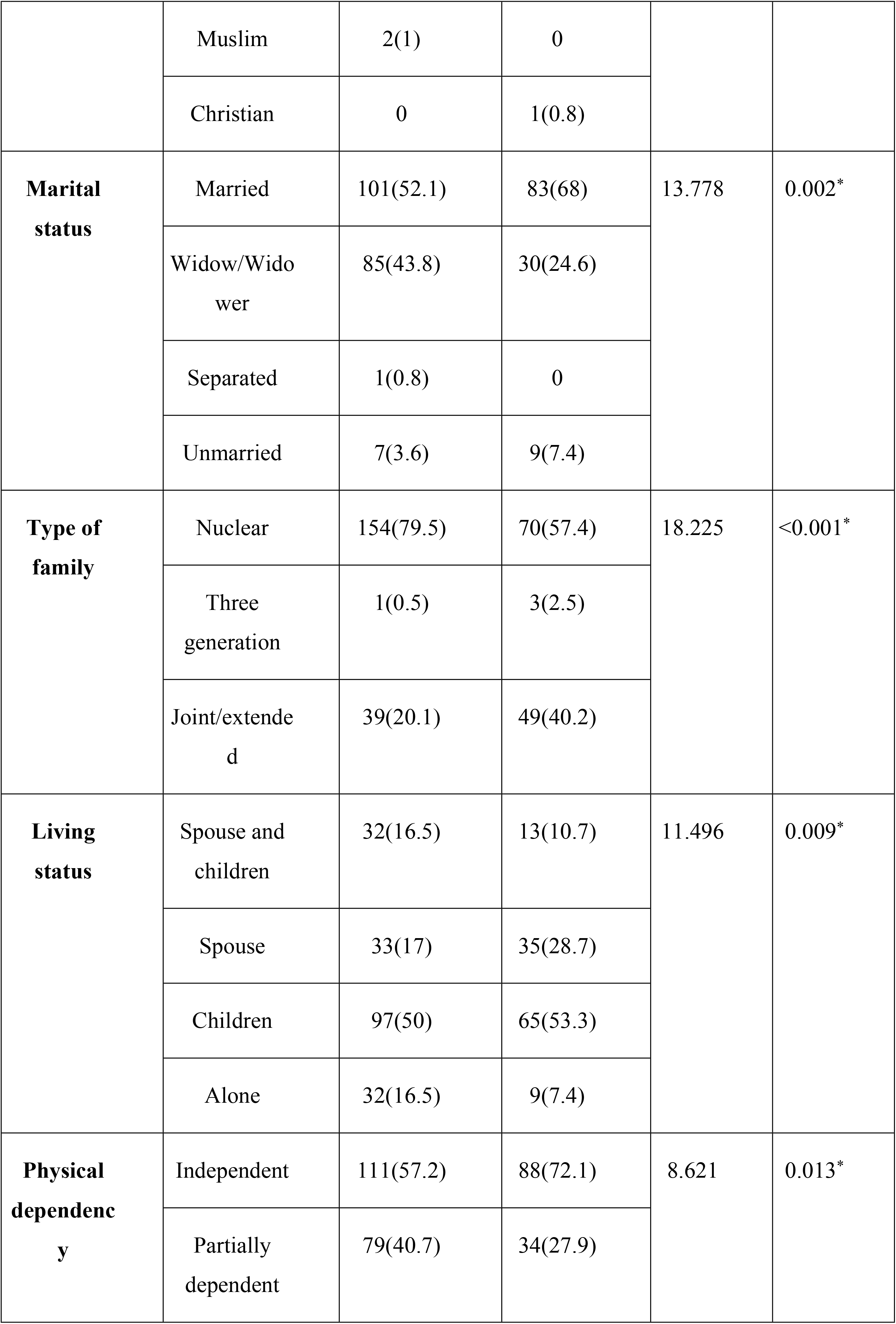
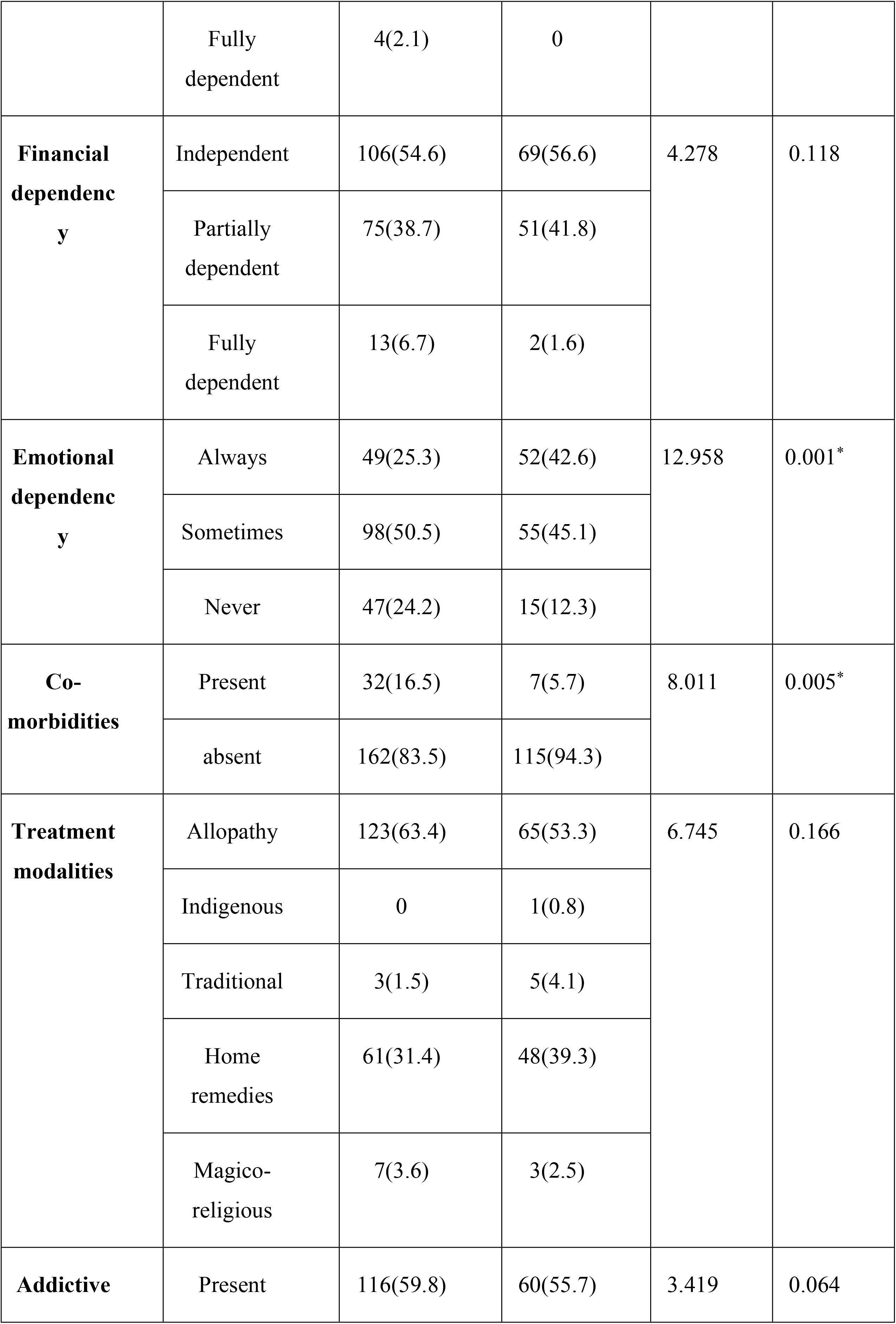
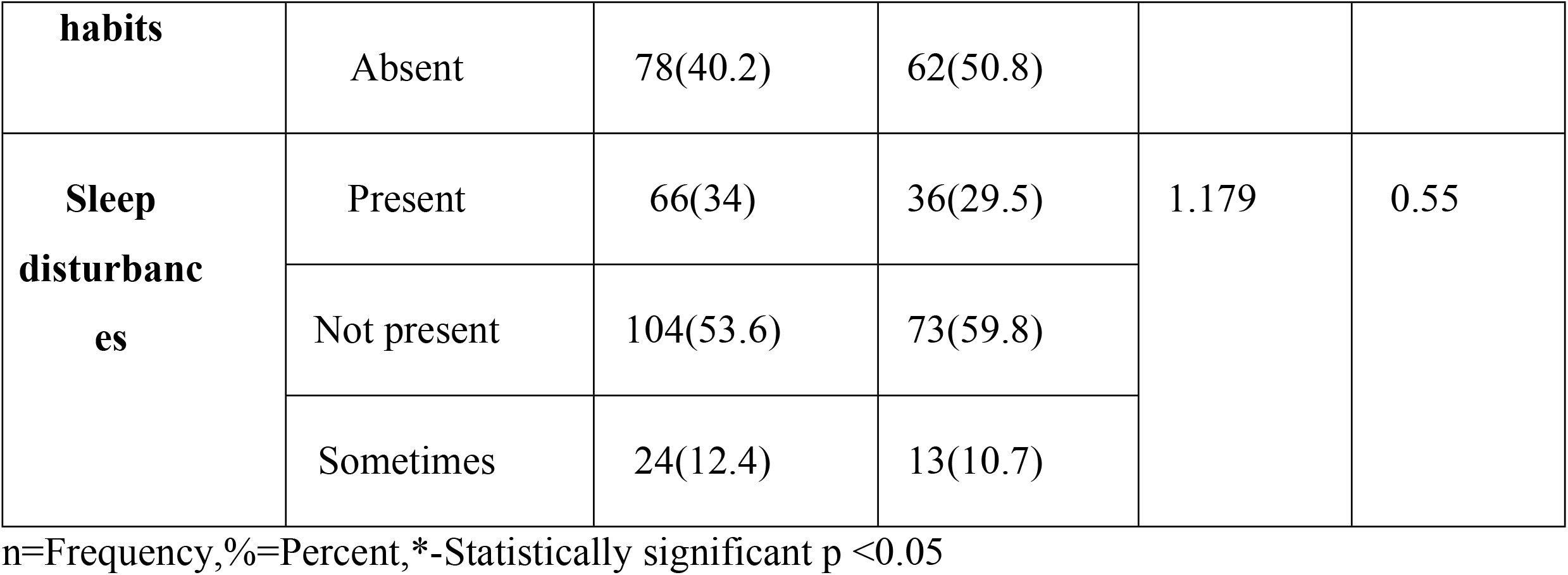
Factors associated with frailty.

### Frailty and cognition

Frailty and cognitive were found to be statistically significant(p<0.05) (table 4). when Pearson correlation was applied it was found that MoCA-B scores and TFI scores are negatively correlated which explained that as cognition increases frailty score decreased but the results were not statistically significant (Fig 4)

**Table 4:**
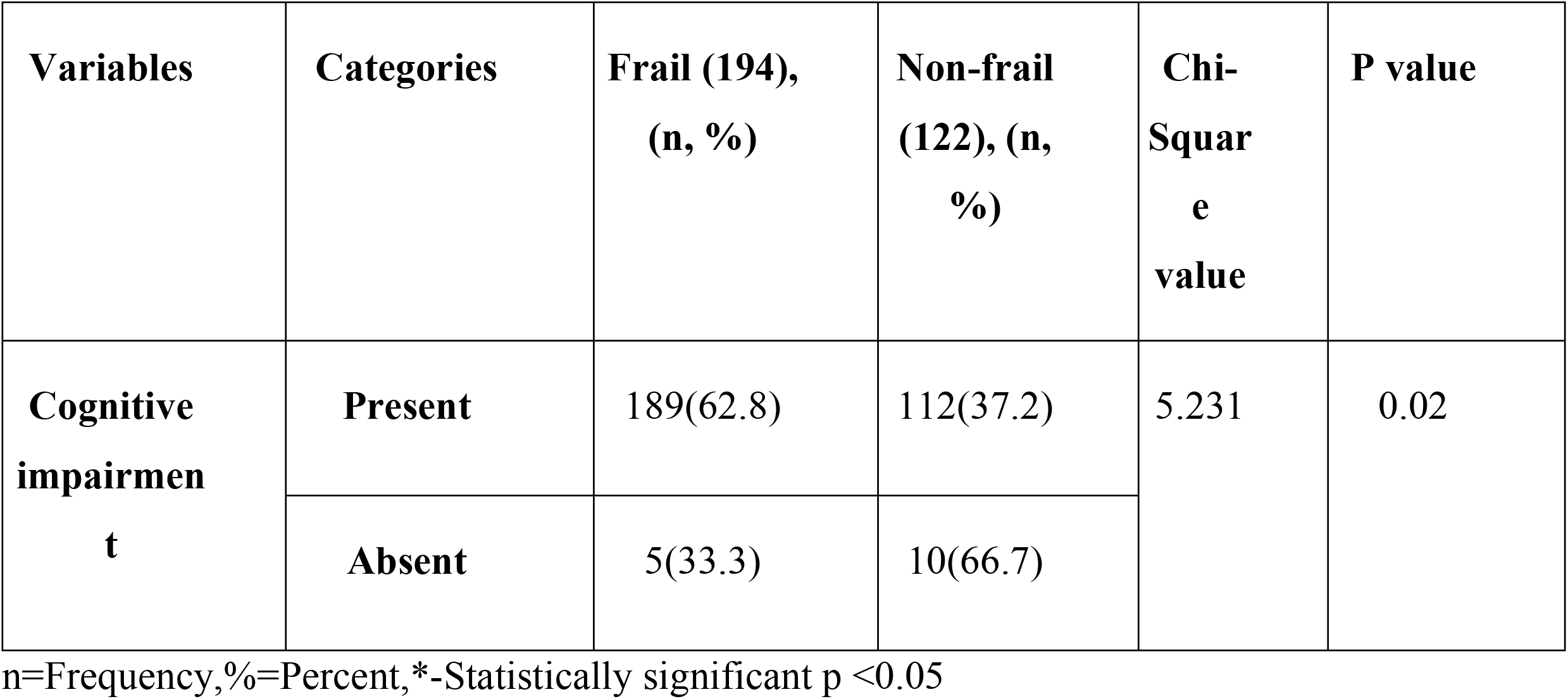
Association between frailty and cognition.

| **Variables** | **Categories** | **Frail (194),**<br>**(n, %)** | **Non-frail**<br>**(122), (n,**<br>**%)** | **Chi-Square**<br>**value** | **P value** |
| --- | --- | --- | --- | --- | --- |
| **Cognitive**<br>**impairment** | **Present** | 189(62.8) | 112(37.2) | 5.231 | 0.02 |
|  | **Absent** | 5(33.3) | 10(66.7) |  |  |

n=Frequency,%=Percent,\*-Statistically significant p <0.05

**Fig 4:**
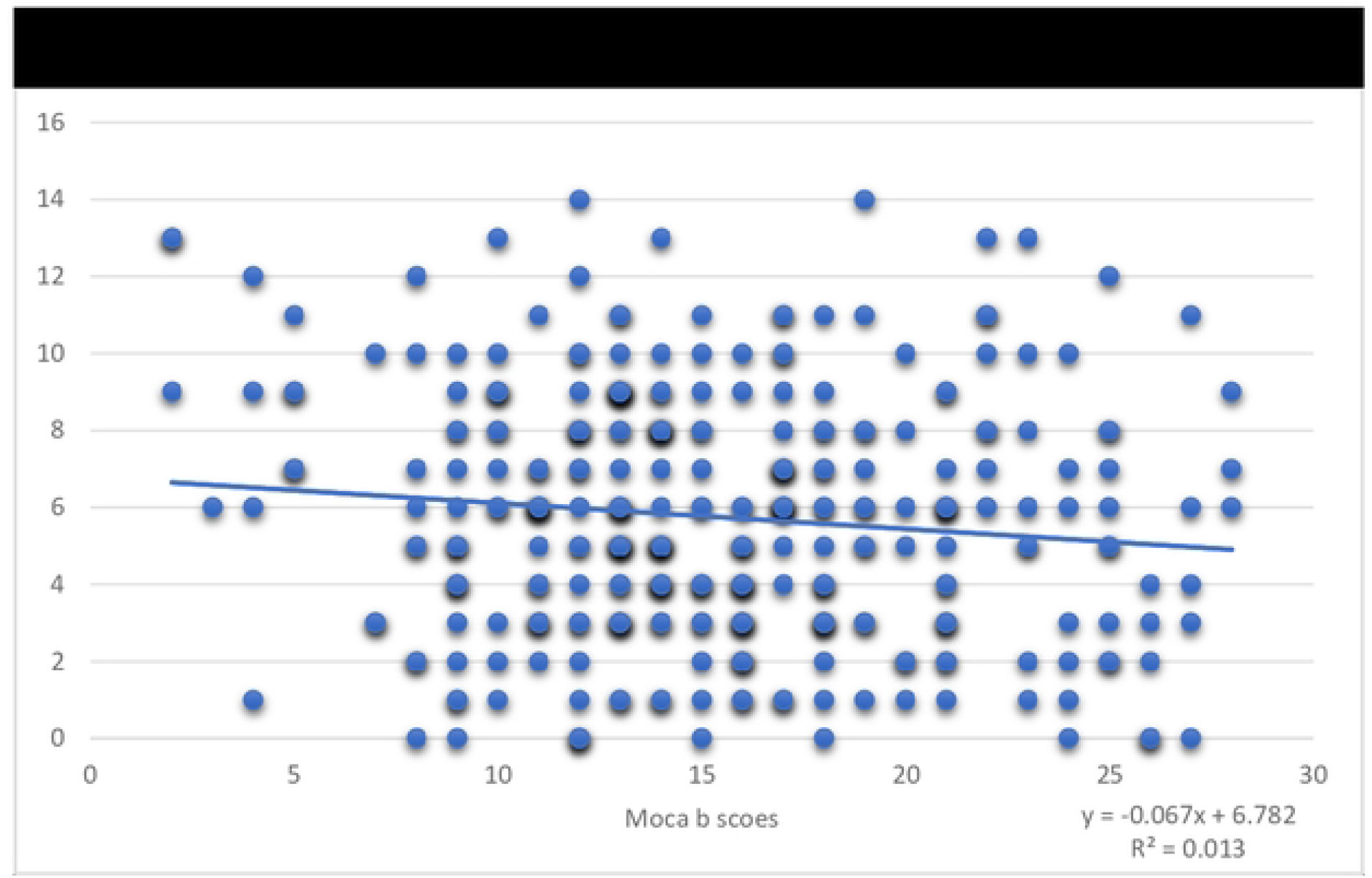
scatter plot which shows the correlation between MoCA-B scores and TFI scores.

## Discussion

Healthy ageing is a significant challenge in this 21^st^ century. The elderly who lives in tribal regions will serve as a link between the traditional tribal way of life and the modernization initiatives being carried out by the government and non-governmental organisations. Being elderly increases their vulnerability to physical and mental health issues ^[17]^ A study was conducted on 952 tribal people in the Vellore districts of South India by S. Sathiyanarayanan et al. The majority in that survey (82.2%) were illiterate and female (59.8%), which was in accordance with the results of this study ^[18]^.

A study using the Bims scale (brief interview for mental state) done among elders in Chengalpattu, Tamilnadu by *Sujitha P et al* found that nearly 44% had mild-moderate cognitive impairment, and 36% had severe cognitive impairment. Only 20% of the respondents had intact cognitive levels ^[19]^where as in our study when graded (according to the moca B scale)13%,42 elders had severe cognitive impairment,54.1%,171 had moderate and 28.2%,89 had mild cognitive impairment.

In the current study, the prevalence of frailty was 61.4%, and female elderly were frailer than male elderly. Similar results were obtained in a study by R P Thakur et al. ^[20]^, where it was found that their frailty was 83.29% more among female elders than male elders. This might be a result of the study’s participants being predominantly female.

The concomitant prevalence of frailty and cognitive changes can greatly predict mortality in the elderly ^[21]^ According to the study conducted by *Isabela Thaís Machado de Jesus et al* among 247 elders in the city of Sao Carlos,87.4% (216) had cognitive impairment, and 84 (38.88%) had some level of frailty (mild, moderate or severe). Whereas in our study out of 301 elders who had cognitive impairment 189 (62.8) was frail. ^[22]^Another study done among 84 elders on a haemodialysis unit by *Jyotish Chalil Gopinathan* et al Cognitive impairment was present in 89.7% of patients and significantly associated with frailty (P < 0.001, Fisher’s exact test) ^[23]^The variations may be due different scales used for assessing cognition and frailty. The limitation of the study includes a smaller sample size and was not able to cover a larger geographical area.

## Conclusion

Thus, the study showed that the tribal elders had a high prevalence of cognitive impairment and the prevalence of cognitive impairment was relatively high among frail people. Although frailty and cognitive decline are part of ageing, its onset and progression can be delayed by appropriate management. Elders diagnosed with cognitive dysfunction should be counselled and regularly monitored. Elderly who are frail should receive counselling on lifestyle improvements including exercise, appropriate nutrition intake, and frequent health checks, among other things, for healthy ageing. All older people with pre-frailty should receive dietary and pharmaceutical supplements of micro and macronutrients. Geriatric clinics may be established at the primary care level to ensure appropriate care and service for the elderly, Additionally, there aren’t enough medical professionals from tribal communities who would be more inclined to work in these places. Medical outreach camps and mobile health clinics can also contribute to bringing health services to remote populations.

## Data Availability

Data will be provided on request

## Acknowledgement

We authors were very thankful to the health facilitators of Vivekananda memorial hospital, sargur, Mysuru for their contribution for conducting the study. we were also grateful to the department of community medicine, JSS medical college for their cooperation

